# Opportunities and limitations of genomics for diagnosing bedaquiline-resistant tuberculosis: an individual isolate meta-analysis

**DOI:** 10.1101/2023.05.04.23289023

**Authors:** Camus Nimmo, Neda Bionghi, Matthew J. Cummings, Rubeshan Perumal, Madeleine Hopson, Shamim Al Jubaer, Allison Wolf, Barun Mathema, Michelle H. Larsen, Max O’Donnell

## Abstract

**Background:** Clinical bedaquiline resistance predominantly involves mutations in *mmpR5* (*Rv0678*). However, *mmpR5* resistance-associated variants (RAVs) have a variable relationship with phenotypic *M. tuberculosis* resistance. We performed a systematic review to (1) assess the maximal sensitivity of sequencing bedaquiline resistance-associated genes and (2) evaluate the association between RAVs and phenotypic resistance, using traditional and machine-based learning techniques.

**Methods:** We screened public databases for articles published until October 2022. Eligible studies performed sequencing of at least *mmpR5* and *atpE* on clinically-sourced *M. tuberculosis* isolates and measured bedaquiline minimum inhibitory concentrations (MICs). We performed genetic analysis for identification of phenotypic resistance and determined the association of RAVs with resistance. Machine-based learning methods were employed to define test characteristics of optimised sets of RAVs, and *mmpR5* mutations were mapped to the protein structure to highlight mechanisms of resistance.

**Results:** Eighteen eligible studies were identified, comprising 975 *M. tuberculosis* isolates containing ≥1 potential RAV (mutation in *mmpR5, atpE, atpB* or *pepQ*), with 201 (20.6%) demonstrating phenotypic bedaquiline resistance. 84/285 (29.5%) resistant isolates had no candidate gene mutation. Sensitivity and positive predictive value of taking an ‘any mutation’ approach was 69% and 14% respectively. Thirteen mutations, all in *mmpR5*, had a significant association with a resistant MIC (adjusted p<0.05). Gradient-boosted machine classifier models for predicting intermediate/resistant and resistant phenotypes both had receiver operator characteristic c-statistics of 0.73. Frameshift mutations clustered in the alpha 1 helix DNA binding domain, and substitutions in the alpha 2 and 3 helix hinge region and in the alpha 4 helix binding domain.

**Discussion:** Sequencing candidate genes is insufficiently sensitive to diagnose clinical bedaquiline resistance, but where identified a limited number of mutations should be assumed to be associated with resistance. Genomic tools are most likely to be effective in combination with rapid phenotypic diagnostics.

## Background

Bedaquiline is a World Health Organization (WHO) group A drug that is included in all multidrug-resistant tuberculosis (MDR-TB) treatment and is a core part of new six-month all oral regimens for MDR-TB alongside pretomanid and linezolid.^1^ It is also included in new clinical trials for short-course treatment of drug-susceptible TB.^2,3^ Bedaquiline was licensed for clinical use in 2012, with pre-clinical lab studies reporting resistance through mutations in the *atpE* gene encoding a subunit of the target ATP pump.^4^ It was not until four years later that the main mechanism of clinical bedaquiline resistance was identified – mutations in the *mmpR5* (*Rv0678*) efflux pump negative repressor, which leads to overexpression of the MmpL5 efflux pump.^5^ Many resistance-associated variants (RAVs) throughout the *mmpR5* gene have now been reported, most of which are individually rare, and have a variable relationship to phenotypic resistance.^6^ In particular, many putative *mmpR5* RAVs are associated with small increases in bedaquiline minimum inhibitory concentration (MIC) which fall near or under the critical concentration for defining resistance.^7^

The lack of a defined genetic hotspot for bedaquiline resistance makes designing a molecular diagnostic that targets a limited number of high confidence mutations, such as Xpert MTB/RIF or XDR cartridges, challenging. If genotypic-phenotypic correlations can be established, then targeted or whole genome sequencing approaches could be used to diagnose bedaquiline resistance. However, to date the WHO mutation catalogue does not contain any variants confidently associated with bedaquiline resistance,^8^ although the number of *M. tuberculosis* isolates eligible for inclusion was limited by the exclusion of heteroresistant mutations (which are common in *mmpR5*) and exclusion of bedaquiline MICs determined using broth microdilution,^9^ as was performed by the CRyPTIC study – by far the largest standardised genotype-phenotype analysis performed to date with over 12,000 isolates.^10^

In this study, we collate all genotypic and phenotypic bedaquiline resistance data reported from clinical isolates up to October 2022, including the over 100 bedaquiline-resistant isolates from the CRyPTIC study, with almost twice the number of isolates with *mmpR5* mutations included of the most recent analysis.^6^ We use a more inclusive method than the WHO Mutation Catalogue, incorporating heteroresistant mutations and broth microdilution plate MICs, to evaluate genotype-phenotype correlations for putative bedaquiline-resistance conferring variants and assess how effective sequencing of resistance-associated genes would be for the identification of bedaquiline resistance. We evaluate the association of individual mutations with resistance using standard statistical as well as machine-based learning methods, and map specific *mmpR5* mutations to the protein structure to highlight mechanisms of bedaquiline resistance.

## Methods

### Search strategy and study selection

The study protocol was developed using the Preferred Reporting Items for Systematic Reviews and Meta-Analyses (PRISMA) guidelines, and was registered in the PROSPERO database (CRD42022346547). We searched for primary studies in electronic databases PubMed, Cochrane, and EMBASE that were published up to October 2022. We conducted manual journal and article searches in the American Journal of Respiratory and Critical Care Medicine and the New England Journal of Medicine. References from studies were also screened for inclusion. Our search strategy included terms “bedaquiline resistance”, “bedaquiline,” “tuberculosis,” “RAVs,” “drug-resistant tuberculosis,” “Rv0678,” “atpE,” “pepQ,” “mmpL5,” “mmpR,” “Rv2535c,” and “Rv1305”. Concepts were exploded to include all Medical Subject Headings (MeSH) subheadings and reveal any further articles.

Inclusion required that a study (1) measure the number of clinical *M. tuberculosis* isolates or patients with bedaquiline resistance via minimum inhibitory concentration (MIC) values with a technique for which a WHO-approved critical concentration (CC) exists (i.e. MGIT or 7H11 agar), or using broth microdilution as was used for the CRyPTIC study and a good evidence base behind the epidemiological cut-off (ECOFF),^11^ and (2) perform sequencing of at least the *mmpR5* and *atpE* genes. Studies that were not in English (or translated into English), had critical concentrations (CCs) not consistent with the WHO CCs^7^, were irrelevant to the study question, used strictly in vitro studies (i.e. *M. tuberculosis* isolates not obtained from patients), were case reports/case series with less than or equal to 3 patients, or for which the full text was unavailable, were excluded. PRISMA was utilised for reporting search results and final studies included in the systematic review. The following definitions were used: MDR-TB was defined as resistance to isoniazid and rifampicin; rifampicin-resistant TB (RR-TB) was defined as resistance to at least rifampicin; clinical *M. tuberculosis* isolates were samples obtained from patients capable of undergoing MIC testing.

### Data extraction and definitions

Two reviewers (NB and MH) independently screened citations utilising a standardised form with pre-defined inclusion and exclusion criteria and extracted data using a standardised data collection form. In cases of disagreement in screening or data abstraction, the disagreement was referred to a third reviewer (MO) to resolve differences and achieve consensus. Duplicate studies of the same cohort were excluded, and if multiple publications included overlapping cohort data, the most comprehensive study was included. We recorded available information on both isolates and patients. Data included MICs for bedaquiline, numbers of isolates or patients with bedaquiline resistance categorised by pre-treatment or during treatment, RAVs identified, and MICs associated with all identified RAVs. Following article retrieval, we applied the US Preventive Services Task Force criteria for evaluation of internal validity of studies to all included studies.^12^ No studies were excluded due to limitations in the validity as assessed by this tool.

Candidate genes for bedaquiline resistance were deemed to be *atpE, mmpR5* (*Rv0678*), *pepQ*, and *atpB*. Potential resistance-associated variants (RAVs) were non-synonymous single nucleotide variants (SNVs), all indels and any mutations 100 base pairs upstream of the promoter. Variants were included if present at ≥5% frequency. Due to the area of technical uncertainty around the CC for bedaquiline, we deemed isolates with MICs greater than the CC to be resistant, those with MICs at the CC to be intermediate, and those with MICs below to be susceptible. CCs were 1μg/mL in MGIT and 0.25μg/mL on 7H11 agar and broth microdilution plate.^7,11^

### Statistical Methods

To calculate the sensitivity of genomic methods for resistance, all mutations in resistant (or intermediate/resistant) isolates that had been routinely phenotyped were incorporated (excluding studies where phenotyping was only performed on isolates with RAVs). To calculate the positive predictive value (PPV) of mutations for resistance, all isolates that were routinely sequenced were included (excluding studies where only phenotypically resistant isolates were sequenced). Methods used to determine resistance for each included study are listed in Supplementary Table 1.

The association of individual variants with resistance, as well as negative predictive value and specificity, was calculated using a dataset of all extracted isolates from included studies (i.e., all those with resistance or mutations in candidate genes) and the curated ‘reuse’ CRyPTIC dataset of 12,288 isolates.^10^ Isolates with >1 mutation across all candidate genes were excluded from the association of individual variants with resistance. We used previously established methods to calculate the odds ratio (OR), likelihood ratio and p-value for the association of putative RAVs with resistance.^13^ We corrected SNVs, but not indels, for the false discovery rate using the Benjamini-Hochberg procedure. Indels are annotated as the nucleotide position followed by ‘_indel’, while SNVs are annotated with amino acid substitution.

### Machine learning models to determine optimal diagnostic RAVs

To identify an optimised set of RAVs that may enhance diagnosis of phenotypic bedaquiline resistance, we performed feature selection using gradient-boosted machine classifier models to identify RAVs most important in predicting phenotypic resistance (mlr, xgboost R packages). For each resistance phenotype (resistant and resistant-intermediate), classifier models were applied to RAVs that occurred with an absolute frequency ≥3. Select model hyperparameters (learning rate [eta], tree depth [max_depth], and number of trees [nround]) were tuned using 10-fold cross validation, with remaining hyperparameters left at default settings. From each classifier model, we identified the top 5 and 10 most important (i.e. predictive) RAVs for each resistance phenotype based on their respective split-gain values. Discrimatory performance of parsimonious RAV sets for each resistance phenotype (dependent variable) was evaluated by generating multivariable logistic regression models that included the top 5 and 10 most important RAVs as independent variables. As several RAVs predicted both resistance phenotypes perfectly (i.e. complete separation was present in the dataset), we applied the likelihood penalty developed by Firth to our logistic models using the logistf R package.^14^ For each Firth-penalised logistic model, we generated receiver-operating-characteristic (ROC) curves and computed the area under each curve (AUC-ROC) with associated 95% confidence intervals (CI), the latter of which were derived using 10,000 bootstrapped replicates.

To minimise classifier model overfitting, we split our sample of isolates into random derivation (70%; N=682) and validation (30%; N=293) sets (*caTools* R package). For each resistance phenotype, predictive RAV sets (i.e. top 5 and 10 most important) were determined exclusively in the derivation set, with discriminatory performance of each set determined in the derivation and validation sets independently.

### Mapping RAVs onto a 3D *mmpR5* protein model

The crystal structure of *mmpR5* was reported by Radhakrishna et al^15^ and deposited as 4NB5 (https://www.rcsb.org/structure/4nb5). The *mmpR5* 4NB5 file was rendered in PyMol (The PyMOL Molecular Graphics System, Version 2.0 Schrödinger, LLC) and bedaquiline RAVs were depicted as stick structures (Figure 3).

## Results

### Predictive value of variants for bedaquiline resistance

Eighteen studies met inclusion criteria (Figure 1, Supplementary Table 1). These studies yielded 975 independent isolates containing ≥1 variant in at least one candidate resistance-associated gene (i.e. potential RAV), of which 545 had ≥1 potential RAV in *mmpR5*. Phenotypic resistance testing demonstrated an intermediate MIC (MIC = CC) for 132 and resistant MIC (MIC > CC) for 201 isolates with candidate gene mutations. An additional 195 intermediate and 84 resistant isolates were identified with no mutations in candidate genes, giving a total of 1254 included isolates of which 274 (21.9%) were fully bedaquiline resistant. 943/1254 (75.2%) were derived from the CRyPTIC study.^10^

**Figure 1.**
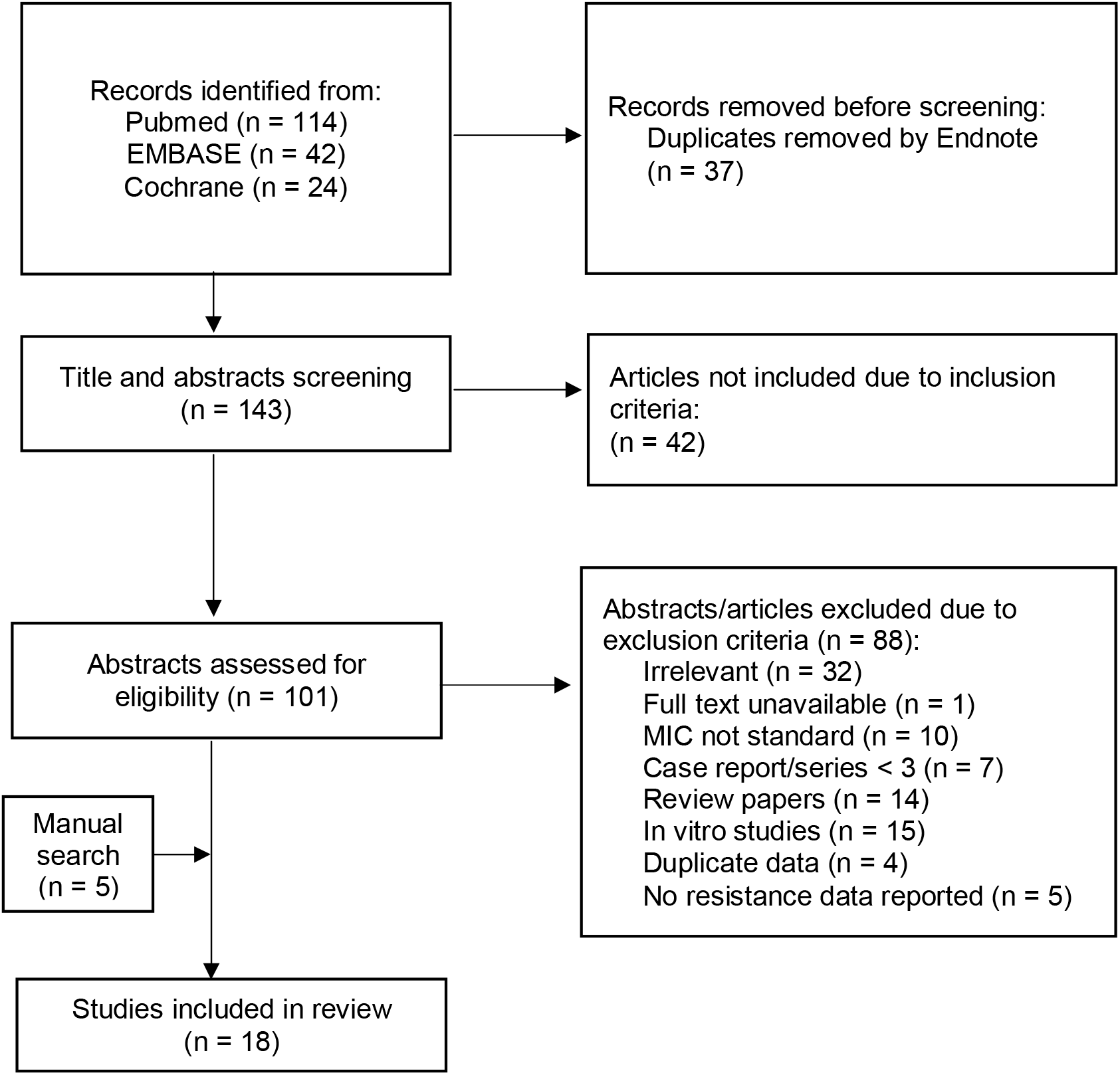
Study profile.

Seven isolates had mutations in >1 candidate gene (Supplementary Table 2).

To determine the maximum achievable sensitivity for phenotypic bedaquiline resistance, our first approach was to assume that any mutation (non-synonymous SNV, promoter mutation or indel) in any candidate gene (*mmpR5, atpE, atpB, pepQ*), or *mmpR5* alone, may cause resistance. Sensitivity of both methods was close to 50% for intermediate/resistant isolates, with inclusion of *atpE, atpB* and *pepQ* increasing sensitivity by 4% (Table 1). Sensitivity for full resistance was higher at over 65% for both methods. Positive predictive value was highest when using any *mmpR5* mutation to predict an intermediate/resistant phenotype at 49%. It was lower for *atpE*, and <10% for *atpB* and *pepQ* genes which contained many variants not associated with resistance (Table 1).

**Table 1.** Sensitivity, positive predictive value, specificity and negative predictive value of different genomic approaches for determination of bedaquiline intermediate/resistant or resistant isolates.

| Any mutation in | Intermediate or resistant | Resistant only |
| --- | --- | --- |
| ***Sensitivity*** |  |  |
| *mmpR5 / atpE / atpB / pepQ* | 310/591 (52.5%) | 190/276 (68.8%) |
| *mmpR5* | 288/591 (48.7%) | 183/276 (66.3%) |
| ***Positive predictive value*** |  |  |
| *mmpR5 / atpE / atpB / pepQ* | 250/891 (28.1%) | 126/891 (14.1%) |
| *mmpR5* | 228/462 (49.4%) | 120/462 (26.0%) |
| *atpB* | 13/210 (6.2%) | 4/210 (1.9%) |
| *atpE* | 9/19 (47.4%) | 7/19 (36.8%) |
| *pepQ* | 7/207 (3.4%) | 1/207 (0.5%) |
| ***Specificity*** |  |  |
| *mmpR5 / atpE / atpB / pepQ* | 11,135/11,777 (94.5%) | 11,330/12,104 (93.6%) |
| *mmpR5* | 11,542/11,777 (97.6%) | 11,753/12,104 (97.1%) |
| ***Negative predictive value*** |  |  |
| *mmpR5 / atpE / atpB / pepQ* | 11,135/11,414 (97.6%) | 11,330/11,414 (99.3%) |
| *mmpR5* | 11,542/11,844 (97.5%) | 11,753/11,844 (99.2%) |

We then evaluated the specificity and negative predictive value of inferring resistance from the methods with sensitivity approaching at least 50%: any mutation in any candidate gene, or from *mmpR5* only (Table 1). We used a simulated highly drug resistant population combining all isolates from included studies and combined this with the entire CRyPTIC ‘reuse’ database (including an additional 11,135 isolates which were bedaquiline susceptible and were wild type in candidate genes). The CRyPTIC dataset includes a significant number of drug-resistant isolates (39% rifampicin-resistant).^16^ Specificity was high (≥94%), as was negative predictive value (>97%), consistent with the fact that bedaquiline resistance is relatively rare in this dataset (<3% with intermediate/resistant phenotype).

### Association of individual variants with resistance

Across all candidate genes there were 62 variants that occurred independent of other candidate gene variants ≥3 times and were selected for further evaluation (Supplementary Tables 3 and 4). Twenty-four variants had a significant association with an intermediate/resistant MIC – 23 in *mmpR5* and one in *atpB* (Table 2, Supplementary Table 3). Most had a strong association (OR >10), except *mmpR5* 192_indel, which had a significant association but weaker strength (OR=6.58). Only one *atpE* variant, E61D, was recorded independently ≥3 times. It was associated with one susceptible, one intermediate and one resistant phenotype, although the association did not meet the level required for statistical significance (Supplementary Table 3).

**Table 2.** Candidate gene variants occurring independently in ≥3 isolates, showing p-value for association with intermediate/resistant and resistant phenotypes. Significant associations are highlighted. Benjamini-Hochberg adjusted p-values for significance was 0.0011.

| Gene | Variant | Intermediate/Resistant phenotype |  | Resistant phenotype |  |
| --- | --- | --- | --- | --- | --- |
|  |  | p-value | Odds ratio | p-value | Odds ratio |
| *mmpR5* | 132_indel | 0.0070 | 38.49 | >0.9999 | 0.00 |
| *mmpR5* | 137_indel | <0.0001 | - | <0.0001 | - |
| *mmpR5* | 138_indel | <0.0001 | 57.73 | <0.0001 | 19.30 |
| *mmpR5* | 139_indel | <0.0001 | - | <0.0001 | 297.29 |
| *mmpR5* | 140_indel | <0.0001 | 57.73 | 0.0890 | 14.16 |
| *mmpR5* | 141_indel | <0.0001 | 41.99 | <0.0001 | 25.10 |
| *mmpR5* | 144_indel | <0.0001 | 211.68 | <0.0001 | 212.35 |
| *mmpR5* | 192_indel | <0.0001 | 6.58 | 0.0290 | 3.61 |
| *mmpR5* | 198_indel | <0.0001 | - | <0.0001 | 127.41 |
| *mmpR5* | 211_indel | 0.0070 | 38.49 | >0.9999 | 0.00 |
| *mmpR5* | 274_indel | <0.0001 | 76.97 | 0.0050 | 28.31 |
| *mmpR5* | 344_indel | <0.0001 | - | <0.0001 | - |
| *mmpR5* | all_del | <0.0001 | 57.73 | 0.0890 | 14.16 |
| *mmpR5* | E21D | <0.0001 | - | 0.0020 | 84.94 |
| *mmpR5* | R50Q | <0.0001 | - | 0.0020 | 84.94 |
| *mmpR5* | Q51R | <0.0001 | - | 0.0670 | 21.24 |
| *mmpR5* | S63R | <0.0001 | - | <0.0001 | 212.35 |
| *mmpR5* | I67S | <0.0001 | - | <0.0001 | - |
| *mmpR5* | N70D | <0.0001 | - | 0.0020 | 84.94 |
| *mmpR5* | R90C | <0.0001 | 48.11 | <0.0001 | 16.99 |
| *mmpR5* | L117R | <0.0001 | 38.49 | 0.1300 | 8.49 |
| *mmpR5* | G121R | <0.0001 | - | <0.0001 | - |
| *mmpR5* | R123K | 0.0010 | 28.87 | 0.1100 | 10.62 |
| *atpB* | T166M | <0.0001 | 12.25 | 0.0630 | 5.31 |

Restricting the analysis to only resistant MICs, 13 variants had a significant association, all in *mmpR5* and all with a strong association except *mmpR5* 192_indel (OR=3.61) (Table 2, Supplementary Table 4).

### Optimised sets of RAVs for prediction of phenotypic resistance

Across all isolates, 71 RAVs that occurred with frequency ≥3 (including occurrences that were not independent of other candidate gene variants) were included in each gradient-boosted machine classifier model. For the intermediate/resistant phenotype, AUC-ROC in the derivation set was 0.73 (95% CI 0.70-0.76) and 0.66 (95% CI 0.64-0.69) for the top 10 and 5 most important RAVs, respectively (Figure 2a). For the resistant phenotype, AUC-ROC in the derivation set was 0.73 (95% CI 0.69-0.76) and 0.66 (95% CI 0.62-0.69) for the top 10 and 5 most important RAVs, respectively (Figure 2b). Ranked importance of RAVs in predicting resistant and resistant/intermediate phenotypes in each classifier model are presented in Figure 2c and d. For both resistance phenotypes, findings were similar in the validation sets (Supplementary Figure 1).

**Figure 2.**
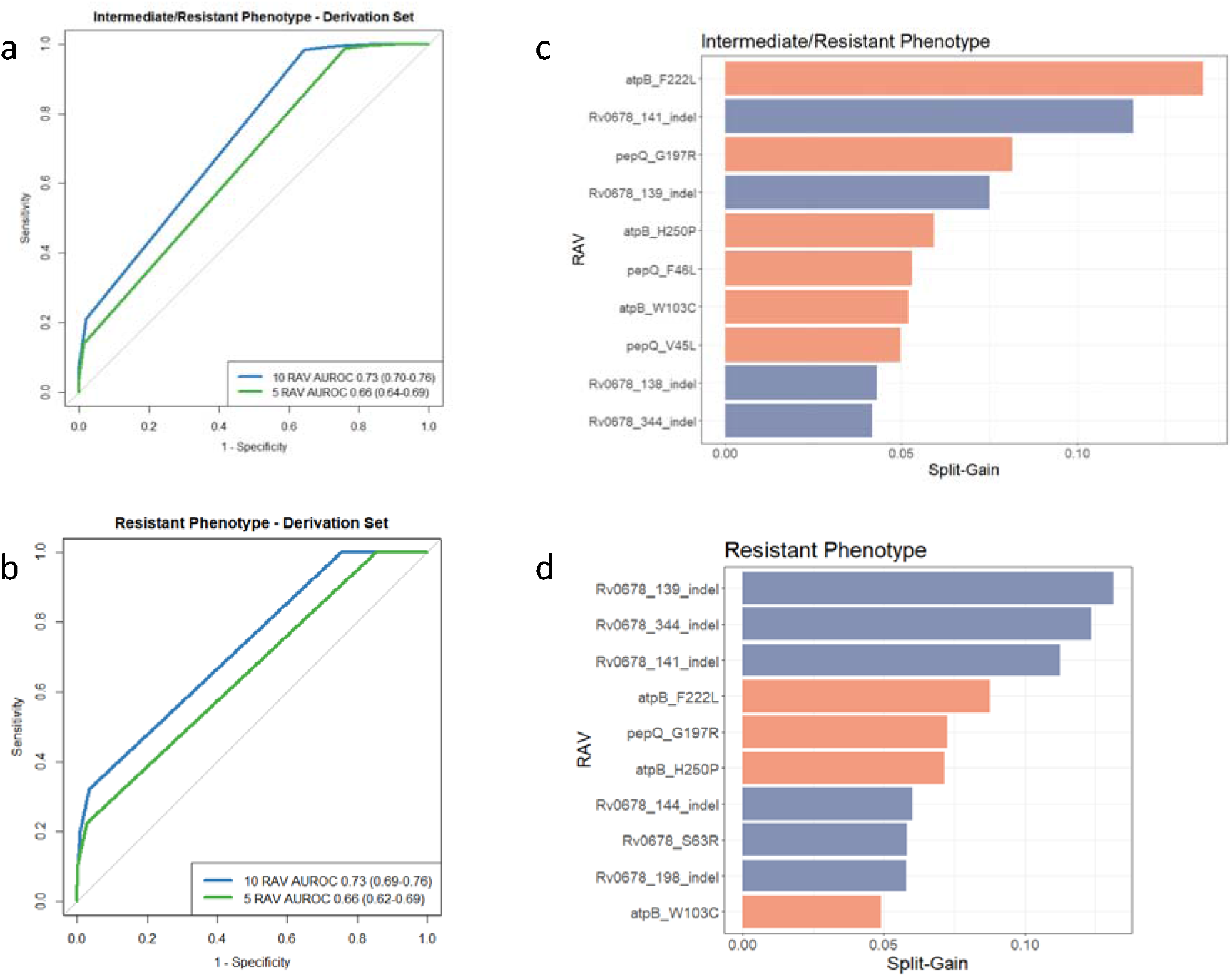
Discriminatory performance of gradient boosted machine classifier models for RAV-based prediction of resistant and resistant/intermediate phenotypes. Receiver operating characteristic (ROC) curves for RAV-based prediction in the derivation cohort for (a) intermediate/resistant phenotype and (b) resistant phenotype. Area under the ROC curve (AUROC) presented with 10,000 bootstrap-derived 95% confidence intervals in parentheses. Top 10 most important RAVs shown for (c) intermediate/resistant phenotype and (d) resistant phenotype. Orange shading indicates association with susceptibility and blue shading with non-susceptibility to bedaquiline.

The mutations identified by machine learning to be most associated with resistance were indels in *mmpR5*, around the hotspot at nucloetides 138-144 as well as positions 198 and 344. These were also strongly associated with resistance in the individual mutation analysis (Table 2). The mmpR5_S63R substitution was also strongly associated with resistance. The remaining 10 most predictive RAVs were predictive of susceptibility rather than intermediate or resistant phenotypes.

### Mapping RAVs onto a 3D mmpR5 protein model

A representation of the *mmpR5* protein structure is shown in Figure 3. Eight of the 14 indels in *mmpR5* associated with bedaquiline resistance map to a 13 base pair stretch (nucleotides 132-144; codons 44-49). These indels are in the coding region for the alpha 2 helix which is part of the DNA binding domain, which correlates with previous reports: frameshift mutations at nucleotides 138-144 and 212-216 were the most frequently associated mutations with bedaquiline resistance that mutations in a 2020 review,^17^ and codons 46-49 and codon 67 were found in 87/199 (43.7%) of South Africa patient isolates demonstrating bedaquiline resistance in a later report.^18^ Such homopolymer indels have been previously implicated in drug resistance in the *glpK* gene.^19^ *mmpR5* has three homopolymer regions of 6 base pairs (nucleotides 139-144), 7 base pairs (nucleotides 192-198), and 5 base pairs (nucleotides 212-216).

**Figure 3.**
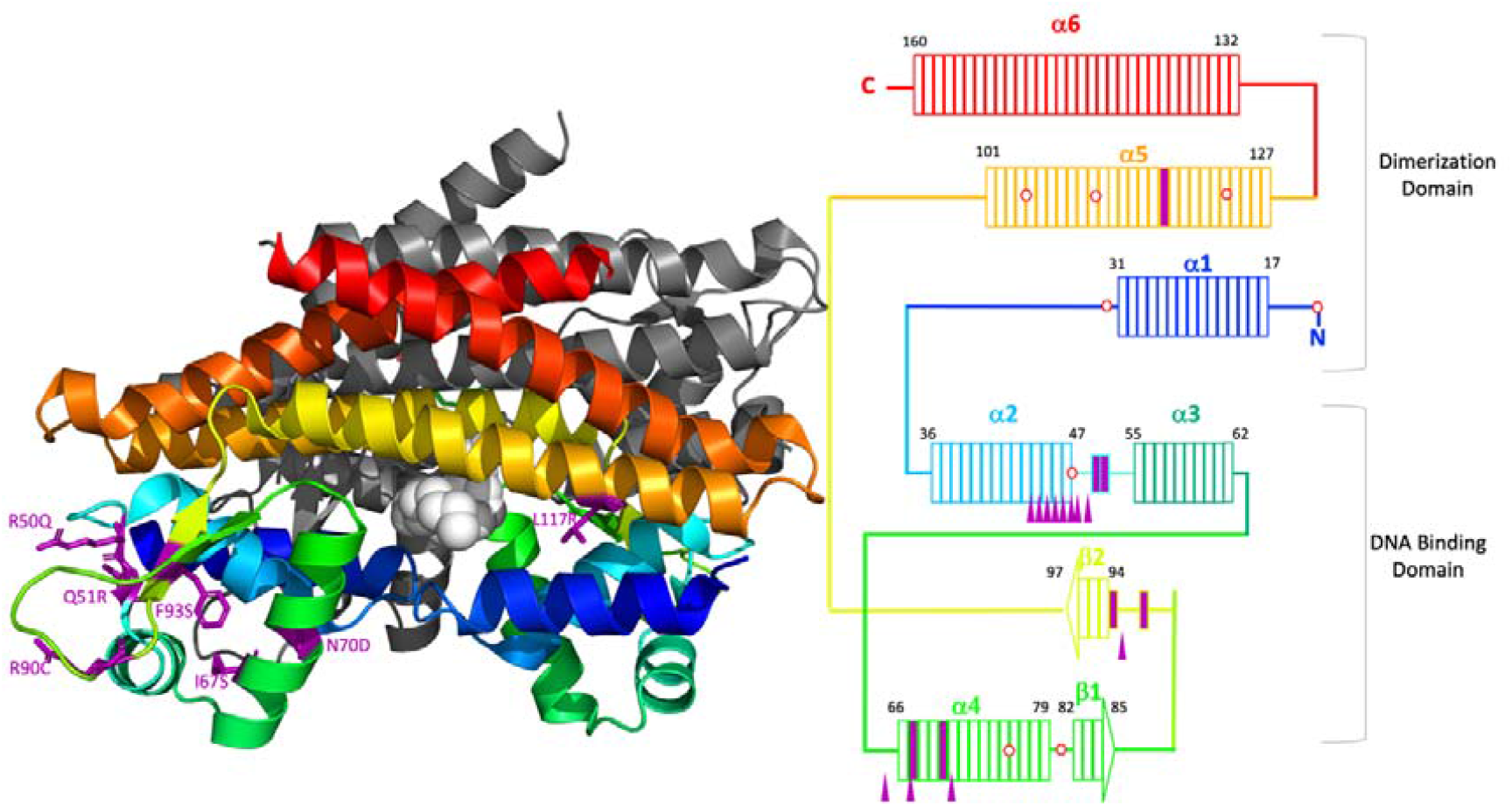
Mapping of bedaquiline resistance associated variants to the secondary structure of *mmpR5* (A) The predicted structure of *mmpR5* (PDB 4NB5) is two homodimers. One homodimer is rendered in colour with a rainbow progression (blue, green, yellow, orange, and red) from the N-terminal with the remaining homodimer in grey. The fatty acid ligand described in Radakhrishan et al15 is rendered in light grey spheres. Amino acid residues from Table 2 that are sites of amino acid substitutions are purple stick structures. (B) Schematic of an *mmpR5* monomer. Rectangles represent amino acids of *mmpR5* and colouring corresponds to the first panel of the figure. The DNA binding domain is comprised of alpha helices 2, 3, and 4 as well as beta sheets 1 and 2. The dimerisation domain is comprised as alpha helices 1, 5, and 6. Amino acid substitutions are indicated by solid purple rectangles. Stop codons in reading frames 2 or 3 are depicted as red hexagons. Indels from Table 1 are shown as purple filled triangles.

DNA binding domain mutations R50Q and Q51R are in the unstructured region between the alpha 2 and alpha 3 helices. At R50Q or Q51R there are substitutions of glutamine and arginine. Comparatively, glutamine is positively charged/hydrophilic and larger than the polar/uncharged arginine. Codons 50 and 51 are in the seven amino acids between alpha 2 and alpha 3 so substitutions of an amino acid with a different size and charge could alter the hinge between these helices which are part of the overall DNA binding domain. The alpha 4 helix is predicted to bind DNA and there are two amino acid substitution mutations I67S and N70D associated with. For I67S the smaller polar amino acid serine with a reactive hydroxyl group is substituted for the larger and very hydrophobic amino acid isoleucine. Also in the alpha 4 helix, an N70D mutation substitutes similarly sized amino acids that with a negatively charged side chain (Aspartic Acid; D) for an amino acid with a polar uncharged side chain (Asparagine; N). There are three loss of function indel mutations in codons 64, 67, and 71. In the eight codons between the beta-1 and beta-2 sheets, this is one loss of function mutation (274_indel) and two amino acid substitutions (R90C and F93S). R90 is a highly conserved residue in the MarR family of transcriptional regulators. The R90C mutation is at a key residue and substitutes the smaller hydrophobic amino acid cysteine for arginine, an amino acid with a positively charged side chain. Aromatic side chains of amino acids can interact with non-protein ligands that contain aromatic groups. These aromatic stacking interactions have been proposed for Y145 and Y157 of *mmpR5*. The F93S amino acid substitution replaces the large and very hydrophobic phenylalanine with an aromatic ring for the much smaller uncharged polar amino acid serine.

The helices alpha-1, alpha-5, and alpha-6 comprise the dimerisation domain. From Table 2, L117R is the only mutation strongly associated with resistance that is found in the dimerisation region. For L117R, the smaller and very hydrophobic leucine is replaced by the much larger hydrophilic arginine that has a reactive side chain.

## Discussion

Even using the most sensitive approach possible, where any mutation in any of the genes that have been identified as potentially associated with bedaquiline resistance is assumed to be resistance-conferring, only around half (49-69%) of bedaquiline resistance would be identified. However, even in this cohort with a high prevalence of drug resistance (41% of the final cohort were rifampicin-resistant), an ‘any mutation’ candidate gene approach would have a high specificity and negative predictive value (>94%), although this is expected given the low rate of overall bedaquiline resistance in the cohort. The remaining phenotype resistance that is unexplained by currently identified genetic targets may be caused by epigenetic mechanisms. *mmpR5* is similar to RND efflux pumps in gram-negative bacteria, which are known to be regulated by both local and global transcriptional regulators, as well as two-component regulatory systems and post-transcriptional modification.^20^

The sensitivity of genomics to predict bedaquiline resistance is therefore less than for the main first-line drugs rifampicin (92%) and isoniazid (90%), and second-line drugs levofloxacin (84%) or moxifloxacin (87%).^8^ It is however equivalent to the sensitivity for pyrazinamide (60%)^8^ which has a similar mechanism of resistance with multiple different loss of function mutations. This is important as to date rapid molecular diagnostic assays have been successfully designed and implemented for rifampicin, isoniazid and fluoroquinolone drugs, although not for pyrazinamide. At sensitivities of 40-60%, genomic tests for pyrazinamide and bedaquiline are both substantially below the 80% sensitivity for detecting resistance required by the WHO’s proposed target product profiles for drug susceptibility testing at peripheral centres.^21^

Although the current understanding of the genetic basis of bedaquiline resistance does not yet lend itself to a standalone rapid diagnostic, whole genome sequencing is already used in several countries such as the UK and USA as the primary resistance test, and current work is underway to develop affordable next generation sequencing that can be used worldwide. Therefore, it is likely that mutations in candidate genes for bedaquiline resistance will be identified, and it is possible that some of these may nevertheless be strongly associated with resistance. Incorporating heteroresistant mutations, using microtitre plate MICs and combining other published studies allowed us to identify 21 mutations that had a strong association with intermediate/resistant bedaquiline MICs, and 11 with resistant bedaquiline MICs. The majority of these were insertions/deletions (65.0% and 61.5% respectively), suggesting that the main mechanism of resistance is loss of gene function. Using machine learning to identify the ten most predictive mutations of phenotype (either susceptible or resistant), several of these mutations (*mmpR5* indels at positions 138-141, 198 and 344) were major contributors. Therefore, where identified by sequencing, these mutations are likely to indicate resistance. Meanwhile, most *pepQ* and *atpB* mutations are unlikely to signify resistance.

Further examining the *mmpR5* protein, most indel RAVs were located in a homopolymer tract in the alpha 2 helix which contains the DNA binding domain. The main substitution RAVs are located in the hinge region between the alpha 2 and 3 helices and the DNA binding region of alpha 4 helix.

A limitation to this study was variability in methodologies used for determining MICs in different settings, although we tried to limit this by including only studies using WHO-approved methods. Also, sequencing varied between sequencing of only candidate genes, and whole genome sequencing. Not all studies sequenced all genes (we required *mmpR5* and *atpE* as a minimum), and their ability to detect low frequency variants differed.

In conclusion, with our current understanding it will remain challenging to adequately diagnose bedaquiline resistance using genomics alone. Given the importance of bedaquiline to RR-TB regimens, establishing adequate access to phenotypic testing must remain a priority. Rapid phenotypic testing methods such as MODS or reporter phage may have a role to play here. However, some variants are strongly associated with resistance, and we would recommend assuming that such isolates are phenotypically resistant pending confirmatory phenotypic testing. Also, due to the high negative predictive value, the absence of resistance-associated variants on target gene sequencing may be interpreted as susceptibility in populations with known low prevalence of phenotypic bedaquiline resistance.

## Data Availability

All data analysed in the manuscript are available online at referenced sources.

