## Supplementary Material for "Opportunities and limitations of genomics for diagnosing bedaquiline-resistant tuberculosis: an individual isolate meta-analysis"

| Study | Primary method(s) of resistance determination | Included for sensitivity analysis | Total isolates | Geno isolates | Pheno isolates | Genes sequenced | MIC method |
| --- | --- | --- | --- | --- | --- | --- | --- |
| Andres 2019^1^ | Phenotype | x | 10 | 10 | 10 | *atpE, mmpR5, mmpS5, mmpL5, pepQ* | MGIT |
| Cryptic Consortium 2022^2^ | Phenotype and genotype | x | 943 | 716 | 353 | WGS | Microtitre |
| Chesov 2022^3^ | Phenotype and genotype | x | 9 | 9 | 9 | WGS | MGIT |
| Conradie 2020^4^ | Phenotype |  | 1 | 1 | 1 | WGS | MGIT |
| Ghodousi 2019^5^ | Phenotype and genotype | x | 8 | 8 | 8 | WGS | MGIT |
| Ismail 2018^6^ | Phenotype and genotype | x | 8 | 6 | 8 | WGS | Microtitre |
| Ismail 2022^7^ | Phenotype | x | 70 | 70 | 69 | WGS | Microtitre |
| Liu 2021^8^ | Phenotype and genotype | x | 11 | 8 | 11 | *atpE, mmpR5, pepQ* | Microtitre |
| Martinez 2018^9^ | Phenotype and genotype | x | 14 | 14 | 3 | WGS | Microtitre |
| Nimmo 2020^10^ | Genotype |  | 21 | 21 | 17 | WGS | 7H11 |
| Saeed 2022^11^ | Phenotype and genotype | x | 20 | 9 | 20 | WGS | Microtitre |
| Veziris 2017^12^ | Phenotype | x | 4 | 3 | 4 | *mmpR5, atpE* | 7H11 |
| Villellas 2017^13^ | Phenotype and genotype | x | 22 | 20 | 11 | *mmpR5, atpE, pepQ* | 7H11 |
| Wu 2021^14^ | Phenotype and genotype | x | 33 | 21 | 27 | *mmpR5, atpE, pepQ* | Microtitre |
| Peretokina 2020^15^ | Phenotype and genotype | x | 23 | 22 | 8 | *mmpR5*, *atpE*, *atpB*, *pepQ* | 7H11 |
| Torrea 2015^16^ | Phenotype and genotype | x | 20 | 19 | 19 | *mmpR5* | MGIT |
| Yang 2018^17^ | Phenotype and genotype | x | 32 | 17 | 28 | *mmpR5*, *atpE* | Microtitre |
| Klopper 2020^18^ | Genotype |  | 1 | 1 | 1 | WGS | MGIT |

Supplementary Table 1. List of studies included and primary method used to determine resistance.

| Source | Gene 1 (mutations) | Gene 2 (mutations) | MIC (method) | Resistance classification |
| --- | --- | --- | --- | --- |
| Peretokina 2020 | *mmpR5* (141_indel) | *atpE* (A63P) | 16 (MGIT) | Resistant |
| Peretokina 2020 | *mmpR5* (138_indel, L114P) | *atpE* (A63V) | 8 (MGIT) | Resistant |
| Peretokina 2020 | *mmpR5* (288_indel) | *atpE* (A63P) | 2 (MGIT) | Resistant |
| Chesov 2022 | *mmpR5* (192_indel) | *atpE* (A63P) | 2 (MGIT) | Resistant |
| Chesov 2022 | *mmpR5* (193_indel, S63G) | *atpE* (E61D, I66M) | 2 (MGIT) | Resistant |
| Cryptic Consortium 2022 | *mmpR5* (141_indel) | *atpB* (T166M) | 1 (microtitre) | Resistant |
| Cryptic Consortium 2022 | *pepQ* (G112D) | *atpB* (W103C) | 0.25 (microtitre) | Intermediate |

Supplementary Table 2. Isolates with variants in >1 candidate gene.

| **Gene** | **Variant** | **Resistant** | **Susceptible/**  **Intermediate** | **p-value** | **Odds ratio** | **LR** |
| --- | --- | --- | --- | --- | --- | --- |
| ***mmpR5*** | 132_indel | 2 | 1 | 0.0070 | 38.49 | 38.61 |
| ***mmpR5*** | 137_indel | 5 | 0 | <0.0001 | - | - |
| ***mmpR5*** | 138_indel | 12 | 4 | <0.0001 | 57.73 | 58.87 |
| ***mmpR5*** | 139_indel | 8 | 0 | <0.0001 | - | - |
| ***mmpR5*** | 140_indel | 3 | 1 | <0.0001 | 57.73 | 58.01 |
| ***mmpR5*** | 141_indel | 24 | 11 | <0.0001 | 41.99 | 43.66 |
| ***mmpR5*** | 144_indel | 11 | 1 | <0.0001 | 211.68 | 215.53 |
| ***mmpR5*** | 192_indel | 13 | 38 | <0.0001 | 6.58 | 6.70 |
| ***mmpR5*** | 198_indel | 8 | 0 | <0.0001 | - | - |
| ***mmpR5*** | 211_indel | 2 | 1 | 0.0070 | 38.49 | 38.61 |
| ***mmpR5*** | 274_indel | 4 | 1 | <0.0001 | 76.97 | 77.47 |
| ***mmpR5*** | 29_indel | 1 | 2 | 0.1410 | 9.62 | 9.64 |
| ***mmpR5*** | 344_indel | 6 | 0 | <0.0001 | - | - |
| ***mmpR5*** | 418_indel | 1 | 5 | 0.2620 | 3.85 | 3.85 |
| ***mmpR5*** | all_del | 3 | 1 | <0.0001 | 57.73 | 58.01 |
| ***mmpR5*** | S2R | 0 | 3 | >0.9999 | 0.00 | 0.00 |
| ***mmpR5*** | N4T | 0 | 6 | >0.9999 | 0.00 | 0.00 |
| ***mmpR5*** | M17V | 0 | 3 | >0.9999 | 0.00 | 0.00 |
| ***mmpR5*** | E21D | 3 | 0 | <0.0001 | - | - |
| ***mmpR5*** | M23V | 0 | 3 | >0.9999 | 0.00 | 0.00 |
| ***mmpR5*** | L40V | 0 | 13 | >0.9999 | 0.00 | 0.00 |
| ***mmpR5*** | R50Q | 3 | 0 | <0.0001 | - | - |
| ***mmpR5*** | Q51R | 3 | 0 | <0.0001 | - | - |
| ***mmpR5*** | E55D | 0 | 13 | >0.9999 | 0.00 | 0.00 |
| ***mmpR5*** | A59V | 1 | 2 | 0.1410 | 9.62 | 9.64 |
| ***mmpR5*** | S63R | 6 | 0 | <0.0001 | - | - |
| ***mmpR5*** | I67S | 3 | 0 | <0.0001 | - | - |
| ***mmpR5*** | N70D | 3 | 0 | <0.0001 | - | - |
| ***mmpR5*** | M73I | 1 | 2 | 0.1410 | 9.62 | 9.64 |
| ***mmpR5*** | I80S | 1 | 3 | 0.1830 | 6.41 | 6.42 |
| ***mmpR5*** | L83P | 2 | 1 | 0.0070 | 38.49 | 38.61 |
| ***mmpR5*** | R90C | 10 | 4 | <0.0001 | 48.11 | 48.89 |
| ***mmpR5*** | F93L | 2 | 1 | 0.0070 | 38.49 | 38.61 |
| ***mmpR5*** | N98D | 1 | 7 | 0.3330 | 2.75 | 2.75 |
| ***mmpR5*** | G103S | 1 | 2 | 0.1410 | 9.62 | 9.64 |
| ***mmpR5*** | L117R | 4 | 2 | <0.0001 | 38.49 | 38.73 |
| ***mmpR5*** | G121R | 3 | 0 | <0.0001 | - | - |
| ***mmpR5*** | R123K | 3 | 2 | 0.0010 | 28.87 | 29.00 |
| ***mmpR5*** | R134G | 1 | 4 | 0.2240 | 4.81 | 4.82 |
| ***mmpR5*** | M139I | 2 | 1 | 0.0070 | 38.49 | 38.61 |
| ***mmpR5*** | L142R | 2 | 1 | 0.0070 | 38.49 | 38.61 |
| ***mmpR5*** | M146T | 2 | 14 | 0.1860 | 2.75 | 2.75 |
| ***atpB*** | G58C | 0 | 5 | >0.9999 | 0.00 | 0.00 |
| ***atpB*** | V87M | 0 | 10 | >0.9999 | 0.00 | 0.00 |
| ***atpB*** | W103C | 0 | 32 | 0.4060 | 0.00 | 0.00 |
| ***atpB*** | T166M | 7 | 11 | <0.0001 | 12.25 | 12.38 |
| ***atpB*** | W216L | 0 | 4 | >0.9999 | 0.00 | 0.00 |
| ***atpB*** | F222L | 0 | 56 | 0.1150 | 0.00 | 0.00 |
| ***atpB*** | H250P | 2 | 46 | >0.9999 | 0.84 | 0.84 |
| ***atpE*** | E61D | 2 | 1 | 0.0070 | 38.49 | 38.61 |
| ***pepQ*** | 818_indel | 2 | 6 | 0.0560 | 6.41 | 6.43 |
| ***pepQ*** | R7Q | 0 | 9 | >0.9999 | 0.00 | 0.00 |
| ***pepQ*** | V45L | 0 | 22 | 0.6250 | 0.00 | 0.00 |
| ***pepQ*** | F46L | 0 | 21 | 0.6230 | 0.00 | 0.00 |
| ***pepQ*** | P69L | 0 | 19 | >0.9999 | 0.00 | 0.00 |
| ***pepQ*** | V104L | 0 | 3 | >0.9999 | 0.00 | 0.00 |
| ***pepQ*** | A124V | 0 | 4 | >0.9999 | 0.00 | 0.00 |
| ***pepQ*** | A187E | 0 | 3 | >0.9999 | 0.00 | 0.00 |
| ***pepQ*** | I193T | 0 | 4 | >0.9999 | 0.00 | 0.00 |
| ***pepQ*** | G197R | 1 | 45 | 0.7280 | 0.43 | 0.43 |
| ***pepQ*** | A242T | 0 | 6 | >0.9999 | 0.00 | 0.00 |
| ***pepQ*** | V328F | 0 | 4 | >0.9999 | 0.00 | 0.00 |

Supplementary Table 3. Associations of variants present ≥3 times in candidate genes with intermediate/resistant phenotype. Significant associations are highlighted. Benjamini-Hochberg adjusted p-values for significance was 0.0011.

| **Gene** | **Variant** | **Resistant** | **Susceptible/**  **Intermediate** | **p-value** | **Odds ratio** | **LR** |
| --- | --- | --- | --- | --- | --- | --- |
| ***mmpR5*** | 132_indel | 0 | 3 | >0.9999 | 0.00 | 0.00 |
| ***mmpR5*** | 137_indel | 5 | 0 | <0.0001 | - | - |
| ***mmpR5*** | 138_indel | 5 | 11 | <0.0001 | 19.30 | 19.63 |
| ***mmpR5*** | 139_indel | 7 | 1 | <0.0001 | 297.29 | 304.75 |
| ***mmpR5*** | 140_indel | 1 | 3 | 0.0890 | 14.16 | 14.20 |
| ***mmpR5*** | 141_indel | 13 | 22 | <0.0001 | 25.10 | 26.25 |
| ***mmpR5*** | 144_indel | 10 | 2 | <0.0001 | 212.35 | 220.04 |
| ***mmpR5*** | 192_indel | 4 | 47 | 0.0290 | 3.61 | 3.65 |
| ***mmpR5*** | 198_indel | 6 | 2 | <0.0001 | 127.41 | 130.13 |
| ***mmpR5*** | 211_indel | 0 | 3 | >0.9999 | 0.00 | 0.00 |
| ***mmpR5*** | 274_indel | 2 | 3 | 0.0050 | 28.31 | 28.51 |
| ***mmpR5*** | 29_indel | 0 | 3 | >0.9999 | 0.00 | 0.00 |
| ***mmpR5*** | 344_indel | 6 | 0 | <0.0001 | - | - |
| ***mmpR5*** | 418_indel | 0 | 6 | >0.9999 | 0.00 | 0.00 |
| ***mmpR5*** | all_del | 1 | 3 | 0.0890 | 14.16 | 14.20 |
| ***mmpR5*** | S2R | 0 | 3 | >0.9999 | 0.00 | 0.00 |
| ***mmpR5*** | N4T | 0 | 6 | >0.9999 | 0.00 | 0.00 |
| ***mmpR5*** | M17V | 0 | 3 | >0.9999 | 0.00 | 0.00 |
| ***mmpR5*** | E21D | 2 | 1 | 0.0020 | 84.94 | 85.53 |
| ***mmpR5*** | M23V | 0 | 3 | >0.9999 | 0.00 | 0.00 |
| ***mmpR5*** | L40V | 0 | 13 | >0.9999 | 0.00 | 0.00 |
| ***mmpR5*** | R50Q | 2 | 1 | 0.0020 | 84.94 | 85.53 |
| ***mmpR5*** | Q51R | 1 | 2 | 0.0670 | 21.24 | 21.31 |
| ***mmpR5*** | E55D | 0 | 13 | >0.9999 | 0.00 | 0.00 |
| ***mmpR5*** | A59V | 1 | 2 | 0.0670 | 21.24 | 21.31 |
| ***mmpR5*** | S63R | 5 | 1 | <0.0001 | 212.35 | 216.13 |
| ***mmpR5*** | I67S | 3 | 0 | <0.0001 | - | - |
| ***mmpR5*** | N70D | 2 | 1 | 0.0020 | 84.94 | 85.53 |
| ***mmpR5*** | M73I | 0 | 3 | >0.9999 | 0.00 | 0.00 |
| ***mmpR5*** | I80S | 0 | 4 | >0.9999 | 0.00 | 0.00 |
| ***mmpR5*** | L83P | 0 | 3 | >0.9999 | 0.00 | 0.00 |
| ***mmpR5*** | R90C | 4 | 10 | <0.0001 | 16.99 | 17.22 |
| ***mmpR5*** | F93L | 0 | 3 | >0.9999 | 0.00 | 0.00 |
| ***mmpR5*** | N98D | 1 | 7 | 0.1700 | 6.07 | 6.09 |
| ***mmpR5*** | G103S | 0 | 3 | >0.9999 | 0.00 | 0.00 |
| ***mmpR5*** | L117R | 1 | 5 | 0.1300 | 8.49 | 8.52 |
| ***mmpR5*** | G121R | 3 | 0 | <0.0001 | - | - |
| ***mmpR5*** | R123K | 1 | 4 | 0.1100 | 10.62 | 10.65 |
| ***mmpR5*** | R134G | 0 | 5 | >0.9999 | 0.00 | 0.00 |
| ***mmpR5*** | M139I | 1 | 2 | 0.0670 | 21.24 | 21.31 |
| ***mmpR5*** | L142R | 1 | 2 | 0.0670 | 21.24 | 21.31 |
| ***mmpR5*** | M146T | 0 | 16 | >0.9999 | 0.00 | 0.00 |
| ***atpB*** | G58C | 0 | 5 | 0.6290 | 0.00 | 0.00 |
| ***atpB*** | V87M | 0 | 10 | >0.9999 | 0.00 | 0.00 |
| ***atpB*** | W103C | 0 | 32 | >0.9999 | 0.00 | 0.00 |
| ***atpB*** | T166M | 2 | 16 | 0.0630 | 5.31 | 5.34 |
| ***atpB*** | W216L | 0 | 4 | >0.9999 | 0.00 | 0.00 |
| ***atpB*** | F222L | 0 | 56 | 0.6410 | 0.00 | 0.00 |
| ***atpB*** | H250P | 0 | 48 | 0.6300 | 0.00 | 0.00 |
| ***atpE*** | E61D | 1 | 2 | 0.0670 | 21.24 | 21.31 |
| ***pepQ*** | 818_indel | 1 | 7 | 0.1700 | 6.07 | 6.09 |
| ***pepQ*** | R7Q | 0 | 9 | >0.9999 | 0.00 | 0.00 |
| ***pepQ*** | V45L | 0 | 22 | >0.9999 | 0.00 | 0.00 |
| ***pepQ*** | F46L | 0 | 21 | >0.9999 | 0.00 | 0.00 |
| ***pepQ*** | P69L | 0 | 19 | >0.9999 | 0.00 | 0.00 |
| ***pepQ*** | V104L | 0 | 3 | >0.9999 | 0.00 | 0.00 |
| ***pepQ*** | A124V | 0 | 4 | >0.9999 | 0.00 | 0.00 |
| ***pepQ*** | A187E | 0 | 3 | >0.9999 | 0.00 | 0.00 |
| ***pepQ*** | I193T | 0 | 4 | >0.9999 | 0.00 | 0.00 |
| ***pepQ*** | G197R | 0 | 46 | 0.6280 | 0.00 | 0.00 |
| ***pepQ*** | A242T | 0 | 6 | >0.9999 | 0.00 | 0.00 |
| ***pepQ*** | V328F | 0 | 4 | >0.9999 | 0.00 | 0.00 |

Supplementary Table 4. Associations of variants present ≥3 times in candidate genes with resistant phenotype. Significant associations are highlighted. Benjamini-Hochberg adjusted p-values for significance was 0.0011. LR = likelihood raito.

| 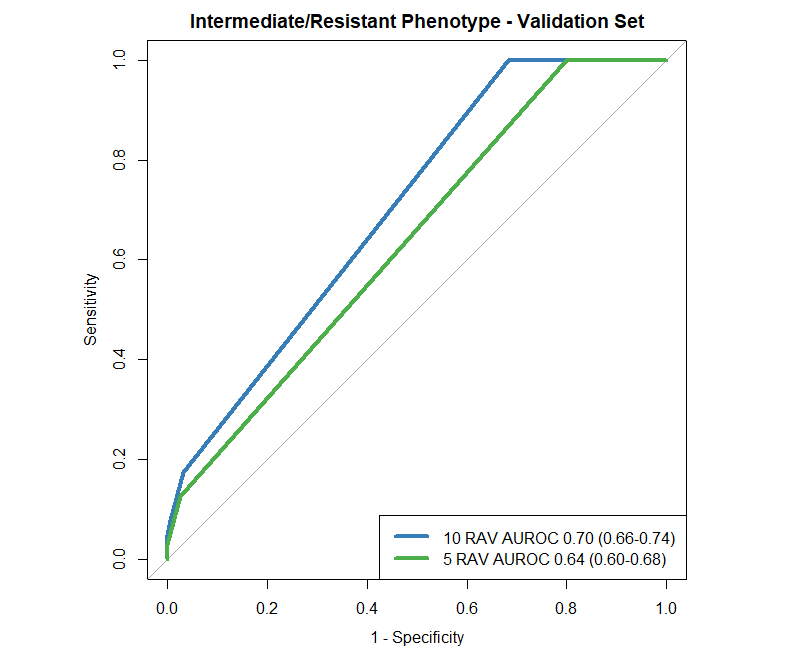 | 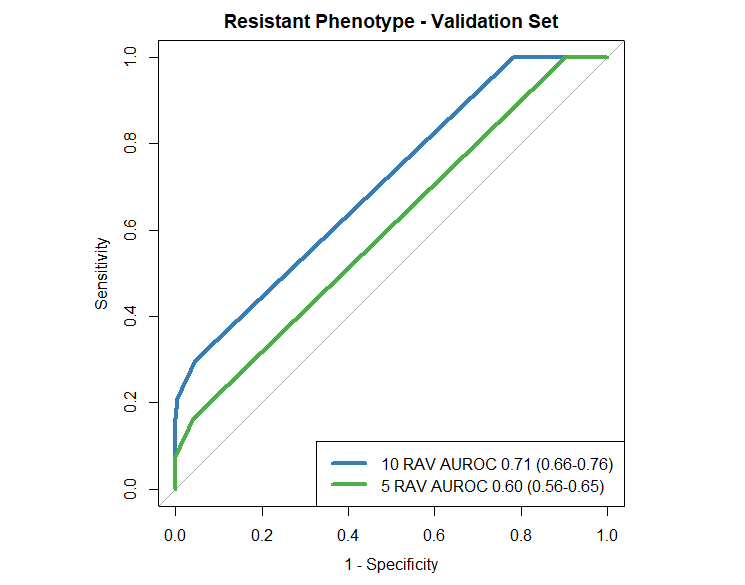 |
| --- | --- |

Supplementary Figure 1. Gradient boosted machine classifier model demonstrating ranked importance of RAVs in predicting resistant and resistant/intermediate phenotypes in each classifier model demonstrating AUC-ROC in the validation cohort for (a) intermediate/resistant phenotype and (b) resistant phenotype.

**Supplementary Material References**

1 Andres S, Merker M, Heyckendorf J, *et al.* Bedaquiline-resistant Tuberculosis: Dark Clouds on the Horizon. *Am J Resp Crit Care* 2019; 0: 1564–8.

2 The CRyPTIC Consortium. A data compendium associating the genomes of 12,289 Mycobacterium tuberculosis isolates with quantitative resistance phenotypes to 13 antibiotics. *Plos Biol* 2022; 20: e3001721.

3 Chesov E, Chesov D, Maurer FP, *et al.* Emergence of bedaquiline resistance in a high tuberculosis burden country. *European Respir J* 2022; 59: 2100621.

4 Conradie F, Diacon AH, Ngubane N, *et al.* Bedaquiline, pretomanid and linezolid for treatment of extensively drug resistant, intolerant or non-responsive multidrug resistant pulmonary tuberculosis. *New Engl J Medicine* 2020; 382: 893–902.

5 Ghodousi A, Rizvi AH, Baloch AQ, *et al.* Acquisition of Cross-Resistance to Bedaquiline and Clofazimine following Treatment for Tuberculosis in Pakistan. *Antimicrob Agents Ch* 2019; 63: e00915-19.

6 Ismail NA, Omar SV, Joseph L, *et al.* Defining Bedaquiline Susceptibility, Resistance, Cross-Resistance and Associated Genetic Determinants: A Retrospective Cohort Study. *Ebiomedicine* 2018; 28: 136–42.

7 Ismail NA, Omar SV, Moultrie H, *et al.* Assessment of epidemiological and genetic characteristics and clinical outcomes of resistance to bedaquiline in patients treated for rifampicin-resistant tuberculosis: a cross-sectional and longitudinal study. *Lancet Infect Dis* 2022; 22: 496–506.

8 Liu Y, Gao M, Du J, *et al.* Reduced Susceptibility of Mycobacterium tuberculosis to Bedaquiline During Antituberculosis Treatment and Its Correlation With Clinical Outcomes in China. *Clin Infect Dis* 2020; 73: e3391–7.

9 Martinez E, Hennessy D, Jelfs P, Crighton T, Chen SC-A, Sintchenko V. Mutations associated with in vitro resistance to bedaquiline in Mycobacterium tuberculosis isolates in Australia. *Tuberculosis* 2018; 111: 31–4.

10 Nimmo C, Millard J, Brien K, *et al.* Bedaquiline resistance in drug-resistant tuberculosis HIV co-infected patients. *Eur Respir J* 2020; 55: 1902383.

11 Saeed DK, Shakoor S, Razzak SA, *et al.* Variants associated with Bedaquiline (BDQ) resistance identified in Rv0678 and efflux pump genes in Mycobacterium tuberculosis isolates from BDQ naïve TB patients in Pakistan. *Bmc Microbiol* 2022; 22: 62.

12 Veziris N, Bernard C, Guglielmetti L, *et al.* Rapid emergence of Mycobacterium tuberculosis bedaquiline resistance: lessons to avoid repeating past errors. *Eur Respir J* 2017; 49: 1601719.

13 Villellas C, Coeck N, Meehan CJ, *et al.* Unexpected high prevalence of resistance-associated Rv0678 variants in MDR-TB patients without documented prior use of clofazimine or bedaquiline. *J Antimicrob Chemoth* 2017; 72: 684–90.

14 Wu S-H, Chan H-H, Hsiao H-C, Jou R. Primary Bedaquiline Resistance Among Cases of Drug-Resistant Tuberculosis in Taiwan. *Front Microbiol* 2021; 12: 754249.

15 Peretokina IV, Krylova LYu, Antonova OV, *et al.* Reduced susceptibility and resistance to bedaquiline in clinical M. tuberculosis isolates. *J Infection* 2020; 80: 527–35.

16 Torrea G, Coeck N, Desmaretz C, *et al.* Bedaquiline susceptibility testing of Mycobacterium tuberculosis in an automated liquid culture system. *J Antimicrob Chemoth* 2015; 70: 2300–5.

17 Yang JS, Kim KJ, Choi H, Lee SH. Delamanid, Bedaquiline, and Linezolid Minimum Inhibitory Concentration Distributions and Resistance-related Gene Mutations in Multidrug-resistant and Extensively Drug-resistant Tuberculosis in Korea. *Ann Lab Med* 2018; 38: 563–8.

18 Klopper M, Heupink TH, Hill-Cawthorne G, *et al.* A landscape of genomic alterations at the root of a near-untreatable tuberculosis epidemic. *Bmc Med* 2020; 18: 24.
